# Relations of Life’s Essential 8 Score with Arterial and Microvascular Function: The Jackson Heart Study

**DOI:** 10.1101/2025.08.01.25332660

**Authors:** Arko S. Dhar, William Hillegass, Leroy L. Cooper, Abu Yusuf Ansari, Ramachandran S. Vasan, Gary Mitchell, Ervin R. Fox

## Abstract

**Background:** Cardiovascular disease (CVD) poses a major burden on the US population, disproportionately impacting African Americans. Vascular function provides a window to assess cumulative risk predisposing individuals to adverse cardiovascular events. The American Heart Association (AHA) Life’s Essential 8 (LE8) provides a means of scoring cardiovascular health, but has yet to be correlated with vascular function.

**Methods:** In a sample of Jackson Heart Study participants (N=2,186, mean age 57 years, 65% women), LE8 scores were calculated per AHA guidelines at baseline visits (2000-2004).

Noninvasive vascular assessments using arterial tonometry and Doppler ultrasound were performed within an ancillary study (2012-2017). Tests measuring aortic and peripheral arterial stiffness included carotid-femoral pulse wave velocity, carotid brachial pulse wave velocity, carotid-radial pulse wave velocity, central pulse pressure, forward pressure wave, and characteristic impedance. Microvascular function tests included baseline and hyperemic brachial artery flow. Linear regression models, adjusted for age, age^2^, sex and heart rate, examined the association between LE8 score (independent variable) and vascular function (dependent variables).

**Results:** In adjusted models, higher LE8 scores were associated with lower carotid-femoral pulse wave velocity (β= -0.32; 95% confidence interval (CI), [-0.42, -0.21]; *p*<0.0001)), characteristic impedance (β= -0.57; 95% CI, [-0.93, -0.20]; *p*=0.0024)), forward pressure wave amplitude (β= - 0.21; 95% CI, [-0.26, -0.16]; *p*<0.0001), central pulse pressure (β= -0.25; 95% CI, [-0.32, -0.19]; *p*<0.0001)) and brachial baseline flow (β= -0.013; 95% CI, [-0.023, -0.002]; *p*=0.021)). Higher LE8 scores were associated with higher brachial hyperemic flow (β= 0.095; 95% CI, [0.035, 0.16]; *p*=0.0018)). When jointly considering the LE8 components in an adjusted model, blood glucose and pressure were most significantly associated with vascular function parameters.

**Conclusions:** Our cross-sectional findings support the concept that a healthy lifestyle is associated with better vascular function. Future longitudinal studies are warranted to investigate whether improving LE8 scores lead to improved vascular function.

**Highlights:**

- A higher AHA Life’s Essential 8 score is significantly associated with lower central artery stiffness and improved microvascular function cross-sectionally.
- Blood glucose and blood pressure are the Life’s Essential 8 components that are significantly associated with change across the most vascular function measures assessed in this study.

## Introduction

Cardiovascular disease (CVD) is a significant health burden for the United States, where approximately 800,000 CVD-related deaths occurred in 2021.^1^ Most CVD events are attributable to modifiable lifestyle behaviors, including diet and level of physical activity, which are associated with the lifetime risk of CVD.^2^ In 2010, the American Heart Association developed the Life’s Simple 7, which was a framework that defined optimal cardiovascular health through the presence of four ideal health behaviors (physical activity, healthy diet, nicotine exposure and normal body mass index) and three health factors (fasting blood glucose, total cholesterol, and blood pressure).^3^ A higher composite Life’s Simple 7 score is associated with a lower risk of all-cause and CVD mortality.^4^

In 2022, the American Heart Association amended the Life’s Simply 7 framework to include sleep hygiene, thereby establishing the Life’s Essential 8 (LE8).^5^ Additionally, the LE8 introduced a broader and more continuous scoring scheme and modified the criteria for multiple existing health factors to better capture widespread variability in individual health behaviors.^5^ Notably, the new LE8 paradigm has a higher predictive value than its predecessor in the setting of adverse CVD events, including myocardial infarction, congestive heart failure, and severe arrythmia, following percutaneous coronary intervention.^6^ A higher LE8 score has also been associated with a decreased risk of incident CVD events in a healthy cohort across multiple age strata.^7^

While previous research has focused on the predictive power of LE8 for CVD events, the putative relations of LE8 score with a comprehensive panel of arterial and microvascular function measures are unknown.^6,7^ Various health factors and behaviors included in the composite LE8 score, including obesity, actively and formerly smoking, elevated blood pressure and high blood glucose and lipid levels are independently associated with disruptions in vascular function.^8^ This study sought to assess associations of composite LE8 and LE8 components (each factor or behavior individually) with arterial and microvascular function measures in a Jackson Heart Study (JHS) sample. It was hypothesized that a lower composite LE8 score, along with lower LE8 component scores, would each be associated with worse arterial and microvascular function.

## Methods

Our cross-sectional study followed the Strengthening the Reporting of Observational Studies in Epidemiology (STROBE) reporting guidelines.^9^ Data from this study was obtained from the Jackson Heart Study, a community-based cohort study assessing risk factors for CVD among African American men and women in Jackson, Mississippi. The procedures for requesting data from the Jackson Heart Study can be found at https://www.jacksonheartstudy.org/.

### Participants

The protocol for the JHS has been previously described.^10,11^ This study’s sample involved a subset of JHS participants who underwent arterial tonometry assessment (2012-2017) and had complete data for LE8 score determination at the first examination cycle (2000-2004) (N=2,186). Participants who did not have sufficient data for LE8 score determination were excluded.

Participants with a history of CVD, including coronary artery disease, stroke, or carotid intervention, were likewise excluded. Figure 1 represents a schematic of the sample selection. Written informed consent was obtained from all study participants, and the study was approved by the Institutional Review Board of the University of Mississippi Medical Center.

**Figure 1:**
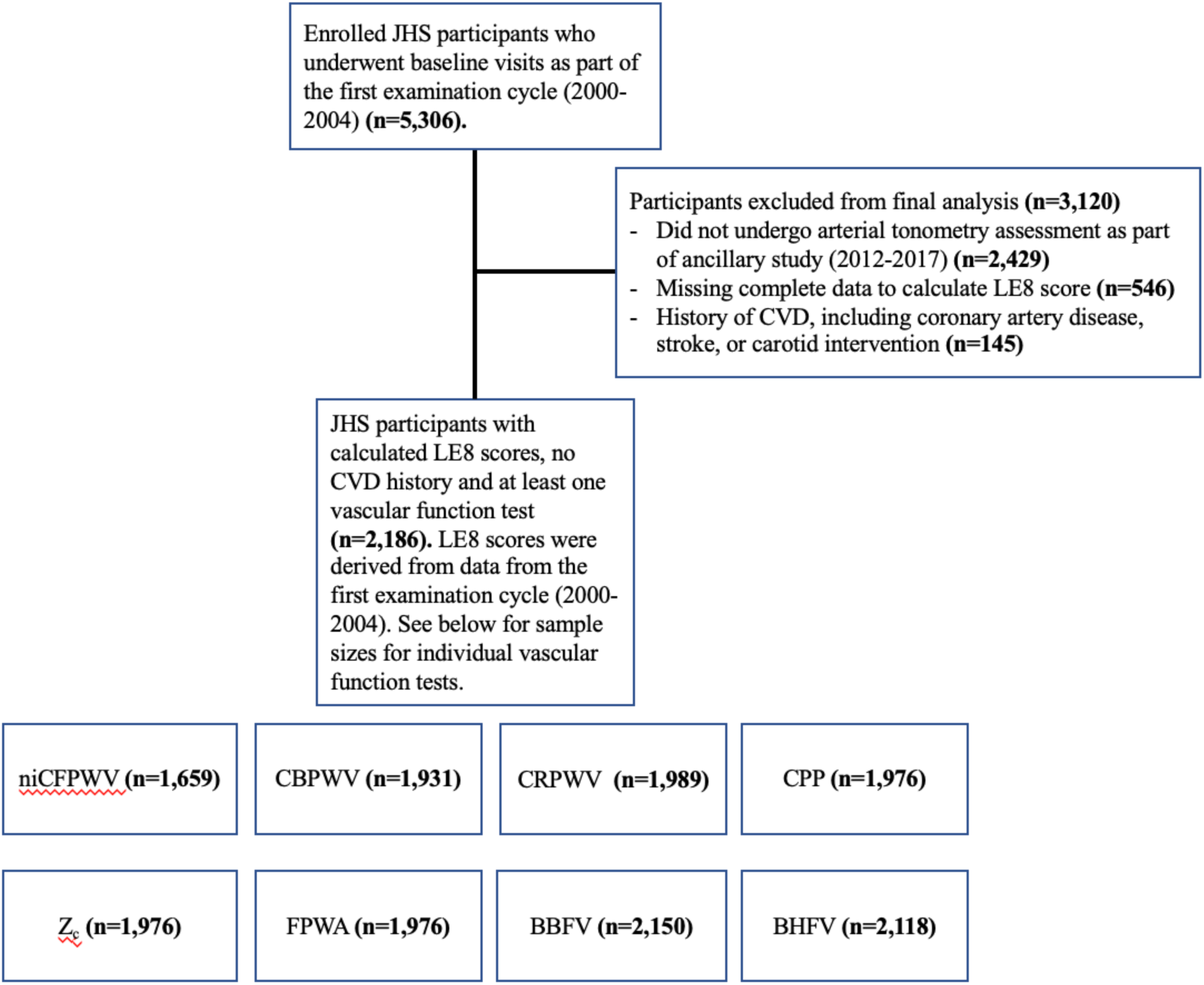
Schematic depiction for inclusion of participants in study analyses. JHS, Jackson Heart Study. LE8, Life’s Essential 8. Vascular function tests include niCFPWV (negative inverse carotid-femoral pulse wave velocity), CPP (central pulse pressure), FPWA (forward pressure wave amplitude), Zc (characteristic impedance), CBPWV (carotid-brachial pulse wave velocity), CRPWV (carotid-radial pulse wave velocity), BBFV (brachial baseline flow velocity) and BHFV (brachial hyperemic flow velocity).

### Hemodynamic assessment with arterial tonometry and ultrasound

Blood pressure was assessed with participants in the supine position following a 5-minute rest, as previously described.^12^ Tonometry was then obtained using simultaneous electrocardiography from the brachial, radial, femoral, and carotid arteries. Two-dimensional images and pulsed Doppler of the left ventricular outflow tract were obtained using echocardiography. Data were subsequently transferred to the core laboratory (Cardiovascular Engineering, Inc, Norwood, MA) for blinded analysis. Tonometry waveforms were signal-averaged, and brachial waveforms were utilized to calculate mean arterial pressure (MAP), per previously described protocols.^12,13^ Diastolic pressures and integrated mean arterial pressure were used to calibrate carotid pressure tracings.^13^ Specific calculations for all vascular function variables were performed as previously described.^14^ Carotid-femoral pulse wave velocity (CFPWV), carotid-brachial pulse wave velocity, and carotid-radial pulse wave velocity were calculated as the ratios of the adjusted transit distance and the pulse transit time difference between the carotid and the femoral, brachial, and radial sites, respectively. The carotid-femoral transit distance was adjusted for parallel transmission as previously described.^15^ Central pulse pressure was calculated as the difference between carotid systolic and diastolic blood pressures. Forward pressure wave amplitude was defined as the difference between the pressure at the foot and at the peak of the forward pressure waveform by performing time domain wave separation analysis using central pressure and flow. Characteristic impedance was calculated in the time domain as the ratio of the pressure increase and the flow increase during the time interval between flow onset and 95% of peak flow, with blood density assumed to be fixed at 1.06 gram/cm^3^ as previously described.^16^

### Microvascular function assessment

Microvascular function was assessed using ultrasound image acquisition and analyses as described previously.^17,18^ Brachial artery Doppler flow was acquired at baseline and following 5 minutes of ischemia produced by inflating a cuff positioned on the forearm. Technicians placed the cuff just distal to the antecubital fold and inflated it to approximately 50 mm Hg above systolic blood pressure. After cuff deflation, sonographers monitored and recorded flow (for 15 seconds after cuff release) until flow peaked. Brachial artery images and Doppler flow were assessed with a Siemens Acuson S2000 ultrasound system mounted with a 9L4 transuducer with a carrier frequency of 9.0 MHz and an insonation angle of approximately 60°. Ultrasound 2D and Doppler audio data were digitized during acquisition and data was transferred to the core laboratory (Cardiovascular Engineering) for blinded analyses. Using a semiautomated signal-averaging technique, flows from the digitized Doppler audio data were analyzed and the timing of peak flow was confirmed from a raw spectral analysis of distinct beats.^19^ 3 to 5 beats (representing the peak flow) were labeled for inclusion in the signal-averaged spectrum. Using the ECG as a fiducial point, flow spectra was signal-averaged and corrected for the actual insonation angle.

### Life’s Essential 8

The American Heart Association LE8 includes four modifiable health behaviors and four biological factors representing an individual’s cardiovascular health status. Specifically, an ideal cardiovascular health involves ≥150 minutes of moderate (or greater) intensity physical activity per week, a Dietary Approaches to Stop Hypertension-style diet score ≥95th percentile, a complete lack of any smoking history including electronic cigarette use, approximately 7-9 hours of sleep per night on average, a body mass index<25 kg/m^2^, non-high-density lipoprotein cholesterol <130 mg/dL, a lack of diabetes mellitus history and free blood glucose <100 mg/dL (or HbA1c <5.7), and a blood pressure <120/<80 mm/Hg.^5^ Per published American Heart Association guidelines, scores on a continuous scale (0-100) were modeled for each component, with an unweighted mean (0-100) representing an individual’s composite LE8 score.^5^ The method by which LE8 scores were derived among JHS study participants by the JHS Coordinating Center is outlined in Tables S1-2.

### Health behaviors under the Life’s Essential 8 score

Self-reported combustible tobacco use data was obtained from participants and categorized as either current (0 points) or never (100 points). Participants who were former tobacco users or ever exposed to secondhand smoking were scored as outlined in Table S1. For dietary score calculation, the Dietary Approaches to Stop Hypertension (DASH) diet score was calculated as previously described.^20^ Components of the dietary score included total fat, saturated fat, protein, fiber, cholesterol, calcium, magnesium and potassium.^20^ Upon calculating a DASH diet score, LE8 score was calculated through comparisons with score percentiles from the National Health and Nutrition Examination Survey during the period of 1999-2004, with the conversions represented in Table S1. For physical activity, the validated Jackson Heart Study Physical Activity Cohort survey was used to gather data from participants.^21^ A score of 100 was given for weekly physical activity greater than 150 minutes, and scores of 0 were given for 0 minutes per week. Intermediate-range values were scored as described in Table S1. For the sleep component of LE8, self-reported hours of sleep were scored as 100 points for 7 to <9 hours, and 0 points for <4 hours. Other durations were scored as outlined in Table S1.

### Health factors under the Life’s Essential 8 score

The non-high density lipoprotein cholesterol of participants was scored as 100 points for <130 mg/dL and 0 points for ≥220 mg/dL, with intermediate-range values being scored as described in Table S2. Body mass index was calculated as weight (kilograms)/height^2^ (meters^2^), with both weight and height being obtained with calibrated devices. Body mass index was scored as 0 points for participants ≥40 kg/m^2^ and 100 points for <25 kg/m^2^, with intermediate values scored as outlined in Table S2. Standard Hawksley random-zero instruments were used to obtain resting, seated blood pressure at 5-minute intervals. Blood pressure was scored as 0 points for systolic pressure ≥160 mm Hg or diastolic pressure ≥100 mm Hg and 100 points for <120/<80 mmHg, with intermediated values being scored as outlined in Table S2. Systolic and diastolic blood pressure values were calibrated using equations provided by the Jackson Heart Study Coordinating Center, as previously described, and these calibrated values were used in the final LE8 score calculation^22^. For blood glucose levels, fasting levels were measured using a Vitros 950 or 250, Ortho-Clinical Diagnostics analyzer (Raritan, NJ). Blood glucose was scored as 0 points for ≥10% glycated hemoglobin and 100 points for participants without self-reported diabetes mellitus and a glycated hemoglobin <5.7 or fasting blood glucose <100 mg/dL. Intermediate blood glucose levels were scored as outlined in Table S2.

### Covariates

Covariates were selected *a priori* as follows: age, age^2^, heart rate and sex. These parameters were all assessed at participants’ baseline visits and are documented in Table 1.

**Table 1:** Sample characteristics among JHS participants (N=2,186) All values are represented as mean±standard deviation or number (%). EGFRckdepi score is used in the assessment of renal function, based on factors including serum creatinine, sex, age and race. “JHS” denotes Jackson Heart Study.

| Variable | Value |
| --- | --- |
| Age (years) | 57±13 |
| Female, N (%) | 1,417 (65) |
| Body mass index, (kg/m<sup>2</sup>) | 32±6.9 |
| Current smoker, N (%) | 218 (10%) |
| Ever smoker, N (%) | 600 (28%) |
| Prevalent hypertension, N (%) | 1092 (50%) |
| Prevalent diabetes, N(%) | 302 (14%) |
| Systolic blood pressure (mm Hg) | 126±15 |
| Diastolic blood pressure (mm Hg) | 76±8 |
| Heart rate (beats/minute) | 66±10 |
| Hemoglobin A1c (%) | 5.8±1.0 |
| Low-density lipoprotein cholesterol, (mg/dL) | 128±36 |
| High-density lipoprotein cholesterol, (mg/dL) | 52±14 |
| Triglycerides (mg/dL) | 104±76 |
| eGFRckdepi score | 97±19 |

### Statistical Analysis

The associations of the composite LE8 score and individual LE8 component scores (independent variables) with vascular function measures (dependent variables) were assessed using multivariable-adjusted linear regression models. Continuous variables were entered as native units in all models. To limit heteroscedasticity, CFPWV was inverted and then multiplied by - 1000 so that higher values corresponded with higher aortic stiffness, which is the conventionally accepted transformation for this measure. A multivariable stepwise analysis was conducted to assess which LE8 components contributed most to the changes in vascular function. Each component score was allowed to be a candidate predictor for each vascular function parameter, with the model ultimately selecting the component scores that were significantly and independently associated with the vascular function result. The threshold p-value for inclusion in the model was p<0.05, and covariates included age, age^2^, heart rate and sex. These were the same covariates used in the models for LE8 composite score versus vascular function, with the threshold p-value for significance in the composite LE8 score model also being p<0.05. To account for multiple comparisons in all statistical models, relationships that were statistically significant under a more conservative Bonferonni-adjusted p<0.00625 were noted.^23^ SAS software (version 9.4, SAS Institute, Cary, NC) to conduct the statistical analysis in this study.

## Results

### Baseline characteristics

The characteristics of study participants are presented in Table 1, along with the LE8 composite score and component score data across our study participants in Table 2. The study sample was comprised of African American adults who were participants in the Jackson Heart Study (N=2,186, mean age 57 years, 65% women).

**Table 2:**
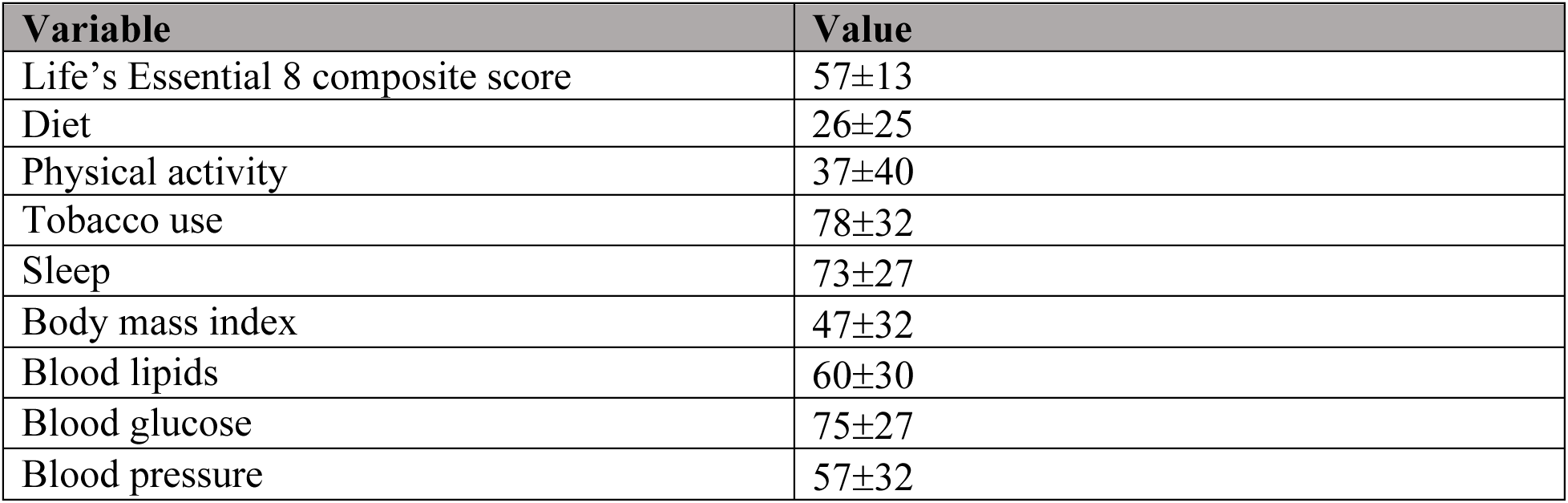
Life’s Essential 8 composite and component scores among JHS participants (N=2,186, mean age 57 years, 65% women) All values are mean ± standard deviation.

| Variable | Value |
| --- | --- |
| Life's Essential 8 composite score | 57±13 |
| Diet | 26±25 |
| Physical activity | 37±40 |
| Tobacco use | 78±32 |
| Sleep | 73±27 |
| Body mass index | 47±32 |
| Blood lipids | 60±30 |
| Blood glucose | 75±27 |
| Blood pressure | 57±32 |

### Associations between composite LE8 score and measures of vascular function

A summary of measures of arterial and microvascular function is presented in Table 3. The associations of composite LE8 score with measures of arterial and microvascular function are presented in Table 4. In linear models adjusted for age, age^2^, heart rate and sex, higher composite LE8 score was associated with lower CFPWV, characteristic impedance, forward pressure wave amplitude, and CPP. Composite LE8 score was not associated with carotid-brachial and carotid-radial pulse wave velocities. Additionally, higher composite LE8 score was associated with higher hyperemic brachial flow velocity but not with baseline brachial flow velocity.

**Table 3:** Mean ± SD of arterial and microvascular function measures among JHS participants. Mean vascular function test results are denoted as mean ± standard deviation (SD). “ni” denotes the inversion and transformation by – 1/1000 that was used to transform CFPWV. CFPWV indicates carotid- femoral pulse wave velocity. “JHS” denotes Jackson Heart Study.

| **Vascular variable** | **Mean ± SD** |
| --- | --- |
| niCFPWV, ms/m (n=1,775) | 11.2±4.47 |
| Central pulse pressure, mm Hg (n=2,105) | 65.4±20.4 |
| Forward pressure wave amplitude, mm Hg (n=2,105) | 53.2±16.0 |
| Characteristic impedance, (dyne·sec)/cm<sup>5</sup> (n=2,105) | 266.3±111.0 |
| Carotid-brachial pulse wave velocity, m/s (n=2,057) | 9.25±2.27 |
| Carotid-radial pulse wave velocity, m/s (n=2,117) | 9.84±1.83 |
| Baseline brachial flow velocity, cm/s (n=2,294) | 5.44±3.16 |
| Hyperemic brachial flow velocity, cm/s (n=2,261) | 47.3±18.8 |

**Table 4:** Associations of composite Life’s Essential 8 score with measures of arterial and microvascular function among JHS participants. Beta regression coefficients (B) – estimated change in outcome variable per 1 unit change in composite LE8 score. Confidence intervals (CI) for the beta coefficients are shown. Models were adjusted for heart rate, age, age^2^ and sex. “ni” denotes the inversion and transformation by – 1/1000 that was used to transform CFPWV. CFPWV indicates carotid-femoral pulse wave velocity. *Denotes association that is statistically significant under the Bonferonni-adjusted threshold (P<0.00625) for multiple comparisons. “JHS” denotes Jackson Heart Study.

| **Vascular variable** | **Est. B (95% CI)** | **P** |
| --- | --- | --- |
| niCFPWV, ms/m (n=1,659) | -0.32 (-0.42, -0.21) | **<0.0001\*** |
| Central pulse pressure, mm Hg (n=1,976) | -0.25 (-0.32, -0.19) | **<0.0001\*** |
| Forward pressure wave amplitude, mm Hg (n=1,976) | -0.21 (-0.26, -0.16) | **<0.0001\*** |
| Characteristic impedance, (dyne·sec)/cm<sup>5</sup> (n=1,976) | -0.57 (-0.93, -0.20) | **0.0024\*** |
| Carotid-brachial pulse wave velocity, m/s (n=1,931) | -0.0065 (-0.015, 0.0016) | 0.11 |
| Carotid-radial pulse wave velocity, m/s (n=1,989) | -0.0039 (-0.010, 0.0026) | 0.24 |
| Baseline brachial flow velocity, cm/s (n=2,150) | -0.013 (-0.023, -0.0019) | **0.02** |
| Hyperemic brachial flow velocity, cm/s (n=2,118) | 0.095 (0.035, 0.15) | **0.0018\*** |

### Multivariable stepwise association of components with measures of vascular function

Tables 5-6 display a multivariable stepwise analysis, which was conducted to identify LE8 components that contributed to relations between LE8 composite score and vascular function measures. For the health behaviors, the diet score was found to be negatively associated with CFPWV and carotid-brachial pulse wave velocity, the physical activity score was found to be negatively associated with CPP, the smoking exposure score was found to be negatively associated with CPP and brachial baseline flow, and sleep was not found to be associated with any of the vascular function tests. For health factors, the blood lipids score was found to be negatively associated with CPP and forward pressure wave amplitude. The blood glucose score was found to be negatively associated with CFPWV, CPP, forward pressure wave amplitude, and characteristic impedance. The blood pressure score was found to be negatively associated with CFPWV, CPP, forward pressure wave amplitude, characteristic impedance, carotid-brachial pulse wave velocity and carotid radial pulse wave velocity. The blood pressure score was positively associated with brachial hyperemic flow. Lastly, the body mass index score was found to be positively associated with characteristic impedance and negatively associated with brachial baseline flow.

**Table 5:** Multivariable adjusted relations between Life’s Essential 8 health behavior component scores with measures of arterial and microvascular function among JHS participants. This table displays all associations that are significant below the p<0.05 threshold. Contributions of the individual LE8 health behavior scores, including diet, PA (physical activity), smoking and sleep, to the vascular function tests, which included niCFPWV (negative inverse carotid-femoral pulse wave velocity), CPP (central pulse pressure), FPWA (forward pressure wave amplitude), Zc (characteristic impedance), CBPWV (carotid-brachial pulse wave velocity), CRPWV (carotid-radial pulse wave velocity), BBFV (brachial baseline flow velocity) and BHFV (brachial hyperemic flow velocity) were assessed. Beta regression coefficients (B) – estimated change in outcome variable per 1 unit change in LE8 component score. Confidence intervals (CI) for the beta coefficients are shown. Models were adjusted for heart rate, age, age^2^ and sex. *Denotes LE8 component contribution to the association that is statistically significant under the Bonferonni-adjusted threshold (P<0.00625). “JHS” denotes Jackson Heart Study.

| **Vascular variable** | **Diet**<br>**Est. B (95% CI)**<br>***P*** | **PA**<br>**Est. B (95% CI)**<br>***P*** | **Smoking**<br>**Est. B (95% CI)**<br>***P*** | **Sleep**<br>**Est. B (95% CI)**<br>***P*** |
| --- | --- | --- | --- | --- |
| niCFPWV<br>(n=1,659) | **-0.067 (-0.12,**<br>**-0.016)**<br>**0.010** | --- | --- | --- |
| CPP (n=1,976) | --- | **-0.024 (-0.043,**<br>**-0.0046)**<br>**0.015** | **-0.035 (-0.060,**<br>**-0.0099)**<br>**0.0062** | --- |
| FPWA(n=1,976) | --- | --- | --- | --- |
| Z<sub>c</sub> (n=1,976) | --- | --- | --- | --- |
| CBPWV<br>(n=1,931) | **-0.0046 (-0.0087,**<br>**-0.00057)**<br>**0.025** | --- | --- | --- |
| CRPWV<br>(n=1,989) | --- | --- | --- | --- |
| BBFV<br>(n=2,150) | --- | --- | **-0.008 (-0.012,**<br>**-0.00379)**<br>**0.0002\*** | --- |
| BHFV<br>(n=2,118) | --- | --- | --- | --- |

**Table 6:** Multivariable adjusted relations between Life’s Essential 8 health factor component scores with measures of arterial and microvascular function among JHS participants. This table displays all associations that are significant below the p<0.05 threshold. Contributions of the individual LE8 health factor scores, including BL (blood lipids), BG (blood glucose), BP (blood pressure), and BMI (body mass index), to the vascular function tests, which included niCFPWV (negative inverse carotid-femoral pulse wave velocity), CPP (central pulse pressure), FPWA (forward pressure wave amplitude), Zc (characteristic impedance), CBPWV (carotid-brachial pulse wave velocity), CRPWV (carotid-radial pulse wave velocity), BBFV (brachial baseline flow velocity) and BHFV (brachial hyperemic flow velocity) were assessed. Beta regression coefficients (B) – estimated change in outcome variable per 1 unit change in LE8 component score. Confidence intervals (CI) for the beta coefficients are shown. Models were adjusted for heart rate, age, age^2^ and sex. *Denotes LE8 component contribution to the association that is statistically significant under the Bonferonni-adjusted threshold (P<0.00625). “JHS” denotes Jackson Heart Study.

| **Vascular variable** | **BL**<br>**Est. B (95% CI)**<br>***P*** | **BG**<br>**Est. B (95% CI)**<br>***P*** | **BP**<br>**Est. B (95% CI)**<br>***P*** | **BMI**<br>**Est. B (95% CI)**<br>***P*** |
| --- | --- | --- | --- | --- |
| niCFPWV (n=1,659) | --- | **-0.11 (-0.16, -0.059)**<br>**<0.0001\*** | **-0.15 (-0.19, -0.11)**<br>**<0.0001\*** | --- |
| CPP (n=1,976) | **-0.040 (-0.066, -0.014)**<br>**0.0027** | **-0.051 (-0.082, -0.021)**<br>**0.001\*** | **-0.098 (-0.12, -0.071)**<br>**<0.0001\*** | --- |
| FPWA(n=1,976) | **-0.027 (-0.048, -0.0066)**<br>**0.01** | **-0.067 (-0.091, -0.042)**<br>**<0.0001\*** | **-0.078 (-0.099, -0.057)**<br>**<0.0001\*** | --- |
| Z<sub>c</sub> (n=1,976) | --- | **-0.40 (-0.58, -0.22)**<br>**<0.0001\*** | **-0.29 (-0.44, -0.14)**<br>**0.0001\*** | **0.19 (0.042, 0.34)**<br>**0.012** |
| CBPWV (n=1,931) | --- | --- | **-0.0042 (-0.0075, -0.00088)**<br>**0.013** | --- |
| CRPWV (n=1,989) | --- | --- | **-0.0033 (-0.0059, -0.00063)**<br>**0.015** | --- |
| BBFV (n=2,150) | --- | --- |  | **-0.0063 (-0.011, -0.002)**<br>**0.004\*** |
| BHFV (n=2,118) | --- | --- | **0.039 (0.015, 0.063)**<br>**0.0015\*** | --- |

## Discussion

This study, which leveraged a large community-based sample of African American individuals in Jackson, Mississippi, demonstrates that American Heart Association’s LE8 modifiable health factors and life behaviors are associated with arterial and microvascular function. Higher LE8 scores were inversely associated with measures of central stiffness, including carotid-femoral pulse wave velocity, central pulse pressure, forward pressure wave amplitude and characteristic impedance, along with being directly associated with measures of microvascular function, namely hyperemic brachial flow velocity (Table 4). LE8 scores were not found to be associated with measures of peripheral artery stiffness, measured through carotid-brachial and carotid-radial pulse wave velocities (Table 4). Our multivariable stepwise model offers insight into LE8 score components that contribute most to relations between LE8 score and variability in measures of arterial and microvascular function, with blood pressure and blood glucose being significantly associated with the largest share of vascular function tests (Table 6). The significant associations in all of our models were present even in the setting of adjusting for multiple traditional risk factors for poor cardiovascular health.

The vascular function tests utilized in this study represent convenient, non-invasive tools for assessing cardiovascular health. Vascular function is a pertinent parameter to investigate as it reflects the global exposure of individuals to various modifiable lifestyle and CVD risk factors, such as diet and exercise, as well as non-modifiable risk factors, such as genetics, age and sex.

The acquired vascular dysfunction phenotypes investigated in this study include diminished microvascular function and large elastic artery stiffening. Hyperemic flow velocity in the brachial artery is influenced by the capacity of distal small vessels to dilate in response to increased metabolic demand, such as after a period of ischemia. This property of small vessels can be compromised by numerous risk factors, including hypertension, diabetes mellitus, oxidative stress, inflammation, and aging.^24–28^ One of the major mechanisms by which these risk factors manifest in microvascular dysfunction is the pathologic remodeling of small vessels.^29–32^ Endothelial dysfunction, while playing a key role in the dilation of the brachial artery in response to ischemia, plays a less significant role in the velocity of blood flow following heightened metabolic demand.^33^ Large artery stiffness, another phenotype assessed in this study, classically manifests in the aorta, which normally serves to damp the pulsatility of pressure in response to blood ejected from the left ventricle. When the aorta stiffens, impedance mismatch between the aorta and muscular arteries is reduced.^17,34,35^ As a result, a larger proportion of a higher level of pulsatile power is transmitted into the conduit arteries and microcirculation. The resulting increased dissipation of pulsatile power in the periphery is associated with microvascular damage and impaired reactivity.^17^ Large artery stiffness shares common risk factors with microvascular dysfunction, and certain risk factors, including elevated blood pressure, glucose and cholesterol, are incorporated into the LE8 score.^5^

While the LE8 score was found to be associated with central arterial stiffness measures, such as CFPWV, it was not found to be associated with peripheral arterial stiffness measures, including carotid-radial and brachial pulse wave velocity. This phenomenon can be explained by the contrasting properties of different types of vessels along the arterial tree. Central arteries, namely the aorta and common carotid arteries, are predominantly composed of elastic fibers, which enables them to expand and buffer increases in stroke volume, protecting vital organs from heavy pulsatile forces.^36^ Peripheral arteries, such as the femoral and brachial arteries, are composed mainly of collagen and smooth muscle, and have a lesser role in capacitance compared to central arteries.^36^ Existing research has elucidated the tendency of central arteries to stiffen with factors including age and the presence of comorbid conditions such as diabetes mellitus and hypertension.^37^ However, increases in peripheral arterial stiffness with age and hypertension have been found to be significantly less pronounced.^38,39^ Of note, in a longitudinal study of patients with renal disease, carotid-radial pulse wave velocity was found to decrease by 0.66 m/s/y, despite an increase in CFPWV of 0.84 m/s/y, a phenomenon that suggests that changes in peripheral arterial composition may not always parallel changes in central arterial composition.^40^ In the setting of this study’s findings, the lack of association of the LE8 score with peripheral arterial stiffness suggests that the LE8 score may not possess the sensitivity to capture changes in peripheral arterial composition. Moreover, measures of central arterial stiffness may carry better value in reflecting the degree of cardiovascular risk conferred by suboptimal LE8 scores.

Existing research has demonstrated an association between the American Heart Association’s LE8 score with cardiovascular disease outcomes, with incremental 10-point increases in LE8 score being linked with significantly lower risk of stroke, myocardial infarction and heart failure across multiple age strata in adjusted models.^7^ Prior research has also demonstrated associations between measures of arterial stiffness and future cardiovascular disease events, with pulse pressure and pulse wave velocity being two parameters shown to be independent predictors of future cardiovascular disease events.^41,42^ In the setting of evident links between both poor vascular function and LE8 score with cardiovascular disease events, the results of this study notably suggest a direct association between LE8 score and vascular function. When assessed alongside existing epidemiological evidence, the association between LE8 and vascular measures supports a plausible paradigm by which vascular dysfunction could mediate the connection between low LE8 score and future cardiovascular disease risk. Further longitudinal and basic-science research is certainly warranted to explore this possibility.

The results of this study not only demonstrate the LE8 score’s association with multiple vascular function tests, but also elucidate which components of the score that serve as the most important determinants of the LE8 relation with vascular function. Importantly, each of the associations assessed in this study are physiologically plausible. For instance, the aggregate adjusted LE8 score was positively associated with brachial hyperemic flow – suggesting that ideal cardiovascular health under LE8 is associated with improved microvascular function (Table 4). Likewise, the adjusted LE8 score was negatively associated with surrogates of central arterial stiffness, including carotid femoral pulse wave velocity and central pulse pressure, among others (Table 4). Thus, as cardiovascular health improves, central arterial stiffness tends to be lower.

With respect to the individual components, it was found that blood glucose and blood pressure were the most influential drivers of the association between LE8 score and vascular function (Table 6). This phenomenon was most pronounced in certain measures of central arterial stiffness, including carotid femoral pulse wave velocity, characteristic impedance, central pulse pressure and forward pressure wave amplitude, where blood pressure and glucose scores were negatively associated with the stiffness parameters (Table 6). The blood pressure score was additionally inversely related with peripheral stiffness measures including carotid-radial and brachial pulse wave velocities, as well as positively associated with a measure of microvascular function, brachial hyperemic flow (Table 6). Taken together, the component score results appear plausible given the significantly deleterious impact that elevated blood pressure and glucose can have on the vasculature.^43,44^ Of note, other LE8 component scores, such as blood lipids, body mass index, diet, physical activity and smoking history, were also negatively associated with certain measures of central arterial stiffness - but not as broadly across the various tests as blood pressure and glucose (Table 5-6). Sleep hygiene, a new component in the LE8 distinguishing the model from the Life’s Simple 7, was not found to be associated with any vascular function test (Table 5). A possible explanation for the close link between elevated blood pressure and glucose and vascular dysfunction is that these risk factors are likely to manifest in cardiovascular complications earlier than others, often when individuals are in a subclinical stage of disease.^45–47^ Existing research has also demonstrated that hypertension and diabetes frequently coexist and have a synergistic relationship, and that they are often common mediators of other factors, such as obesity and sedentary lifestyle, in triggering cardiovascular disease.^48,49^

The strengths of this study include the use of a large cohort of African American individuals living in the US who have been extensively phenotyped at a cardio-metabolic level, including extensive vascular function testing and scoring for ideal cardiovascular health metrics. African Americans have a disproportionately higher risk of experiencing cardiovascular disease, which manifests earlier in life and may become refractory to conventional treatment.^50^ Previous studies have shown that African Americans have reduced microvascular function, measured as reactive hyperemia index (RHI), compared to White individuals.^51^ They were also noted to have greater arterial stiffness, as measured by higher pulse wave velocity.^51^ Strikingly, these differences persisted even in subgroups of individuals completely free of conventional cardiovascular disease risk factors, such as obesity, hypertension, type 2 diabetes mellitus, and tobacco use.^51^ Another strength of our study is that the final results are in line with prior findings from the landmark Framingham Heart Study, which found that correlates of abnormal microvascular function include elevated systolic blood pressure, diabetes status, blood lipids and smoking, among others.^52^ Prior research from the JHS also determined a significant association between risk factors such as elevated blood pressure, glucose and cholesterol, with measures of arterial stiffness.^53^

Of note, the baseline characteristics of our study’s population closely align with the broader US population. The prevalence of diabetes (14% vs. 14%), hypertension (50% vs. 49.6%) and tobacco smoking history (10% vs. 11.6%) are similar.^54–56^ The mean body mass index of our study sample (31.6 kg/m^2^±6.9) is only slightly higher relative to the US population aged 20 and above, 29.1 kg/m^2^.^57^ The exclusion of participants with CVD history is another strength of our study given that it allows for an assessment of the relations of composite LE8 score and the various component measures with subclinical patterns of vascular dysfunction prior to the manifestation of overt cardiovascular disease.

Our study has multiple limitations. Firstly, the time interval between the subjects’ vascular function tests and baseline LE8 score calculations in our participants precludes the determination of a pure cross-sectional association. However, it can be argued that the time interval sheds light on whether LE8 scores are associated with vascular function results after a period of time in which participants are exposed to a level of risk factors to at least the level known in their baseline visits. For the smoking component of the LE8 score, our study methodology assessed history of tobacco (combustible cigarette) use, but failed to account for other forms of smoking such as electronic cigarettes, which also contain nicotine. Another limitation of our study is that while our final analysis did exclude participants with coronary artery disease, stroke and carotid intervention, these exclusion parameters are certainly not inclusive of every possible form of cardiovascular disease. Likewise, while our study does adjust for numerous cardiovascular risk factors, such as age, age^2^, heart rate and sex, it is certainly not inclusive of every possible risk factor that could influence vascular function. From a demographic standpoint, all participants in our study identify as African American, meaning that our findings may not be generalizable to other ethnic or racial groups. Similarly, as our sample comprised mostly middle-aged participants (mean age 57±13), the findings may not be generalizable to younger individuals, or the US population at large, which has a median age of 38.5 years.^58^

## Conclusions

The American Heart Association Life’s Essential 8 was found to be associated with measures of vascular function, including those of large central arteries and the microvasculature in this large, community-based cohort study of African American participants. Composite LE8 scores were associated with all vascular function tests aside from those of the peripheral vessels. Specific components of the LE8 score were also determined to be associated with vascular function measures, with blood pressure and glucose scores being significantly associated with the largest share of vascular function tests. Sleep, a component introduced to the LE8 from the previous Life’s Simple 7, was not associated with any vascular function test. The results of this study add to the growing body of evidence demonstrating that unhealthy lifestyle behaviors, such as smoking and minimal exercise, and health factors, such as high blood pressure, glucose, and cholesterol, are associated with adverse cardiovascular manifestations. The association between the LE8 score and measures of vascular function may help explain the basis behind a previously determined association between poor LE8 score and cardiovascular disease events.^7^ Future longitudinal studies are warranted to further elucidate this complex, multifactorial relationship, and assess how the LE8 score could potentially predict changes in cardiovascular health status overtime.

## Data Availability

All relevant data is included within the manuscript and statistical data files/code are available upon request.

https://pubmed.ncbi.nlm.nih.gov/15367870/

https://pubmed.ncbi.nlm.nih.gov/16320381/

## Acknowledgements

The authors would like to express gratitude to the participants and staff of the Jackson Heart Study.

## Supplemental Material

Tables S1–S5 Reference #5, #20

## Non-standard abbreviations and acronyms

CVD: cardiovascular disease
CFPWV: carotid-femoral pulse wave velocity
CPP: central pulse pressure
LE8: Life’s Essential 8

